# Systematic Modality Ablation of Multimodal Machine Learning for Predicting 24-Month Progression from Mild Cognitive Impairment to Alzheimer’s Disease

**DOI:** 10.64898/2026.09.01.26360413

**Authors:** Sophie Choe

**Affiliations:** Clinovia.ai

**Author notes:** Corresponding author: Sophie Choe, Clinovia.ai.

**Keywords:** Alzheimer’s disease, mild cognitive impairment, disease progression, multimodal biomarkers, biomarker ablation, machine learning, risk prediction, cognitive assessment, magnetic resonance imaging, cerebrospinal fluid biomarkers, positron emission tomography, APOE genotype, XGBoost, clinical utility

## Abstract

**INTRODUCTION:** Multimodal biomarkers have transformed Alzheimer’s disease research, but the incremental contribution of each modality to predicting progression from mild cognitive impairment (MCI) remains unclear. We systematically evaluated individual biomarker modalities using a comprehensive ablation framework.

**METHODS:** We analyzed 2,430 participants with MCI from the Alzheimer’s Disease Neuroimaging Initiative with known 24-month progression status. XGBoost models were trained using combinations of demographic variables, cognitive assessments, apolipoprotein E (APOE) genotype, structural MRI, cerebrospinal fluid (CSF) biomarkers, and PET biomarkers. Performance was evaluated using repeated stratified 5×10 cross-validation; discrimination was assessed using the area under the receiver operating characteristic curve (AUC), with pairwise comparisons via DeLong’s test on single-pass out-of-fold predictions, Holm-Bonferroni corrected. Complete-case analyses evaluated the impact of missing data and median imputation.

**RESULTS:** The full multimodal model achieved the highest discrimination (AUC=0.934). Excluding cognitive assessments produced the largest reduction (AUC=0.883, P<0.001 vs. multimodal), the only comparison to remain significant after correction. Removing APOE, CSF, or MRI produced only modest, statistically indistinguishable reductions (AUC=0.933, 0.931, 0.932). Removing PET produced a similarly small reduction in the primary analysis (AUC=0.932), but complete-case sensitivity analysis showed imputation significantly inflated this estimate (P=0.005); PET’s true contribution may exceed the other three. The baseline clinical model performed near chance (AUC=0.556).

**DISCUSSION:** Cognitive assessment contributes substantially more predictive information for 24-month progression than structural imaging, molecular biomarkers, or genetic risk, whose removal produces only modest, largely statistically indistinguishable performance loss — with the exception of PET, whose apparent equivalence may be an imputation artifact. These findings establish an evidence-based hierarchy of biomarker utility and provide a quantitative framework for designing cost-effective prediction models and prioritizing biomarker acquisition in clinical research and multimodal AI systems.

## Background

Alzheimer’s disease (AD) is a progressive neurodegenerative disorder, and identifying individuals with mild cognitive impairment (MCI) [1,2] who are most likely to progress to AD is an important goal in clinical research and practice. The clinical course of MCI is heterogeneous, with substantial variation in the likelihood and timing of progression to dementia [3]. The emergence of disease-modifying therapies for early AD has further increased the importance of identifying individuals at elevated risk of progression to support clinical decision-making, treatment planning, and clinical trial enrollment [4,5].

A broad range of clinical and biomarker measurements has been developed to characterize the biological and cognitive processes associated with AD. Cognitive assessments are widely used clinical measures that provide direct information about cognitive impairment [6–8]. Structural magnetic resonance imaging (MRI) provides measures associated with neurodegeneration [9], while cerebrospinal fluid (CSF) biomarkers provide information related to amyloid and tau pathology [10,11]. Amyloid and fluorodeoxyglucose (FDG) positron emission tomography (PET) provide complementary information regarding amyloid deposition and cerebral glucose metabolism [12,13]. Apolipoprotein E (APOE) ε4 genotype provides information regarding genetic susceptibility to AD [14]. Together, these modalities capture complementary and partially overlapping aspects of the clinical and biological processes associated with AD, consistent with contemporary biomarker frameworks [15,16].

Machine-learning approaches provide an opportunity to integrate heterogeneous clinical and biomarker measurements and model potentially nonlinear relationships among predictors. Recent studies have therefore increasingly developed multimodal prediction models combining cognitive assessments with neuroimaging, fluid biomarkers, genetic information, and other clinical variables [17–20]. Although integration of multiple modalities can improve predictive performance, high-dimensional multimodal models also introduce substantial practical challenges, including increased data requirements, missingness, cost, invasiveness, and limited availability of specialized biomarker measurements.

An important unresolved question is therefore whether every modality included in a multimodal prediction model provides meaningful incremental predictive information. High predictive performance of an integrated model does not by itself establish that each constituent modality contributes substantially to that performance. A modality may be biologically informative yet provide little additional discrimination once other correlated modalities are available. Conversely, removal of a modality that contains complementary information should produce a measurable reduction in predictive performance. Distinguishing these situations is important for both computational model design and clinical translation.

In a previous analysis of this ADNI cohort [21], we systematically compared individual biomarker modalities under a common machine-learning framework and found that cognitive assessment provided substantially greater discrimination for 24-month MCI-to-AD progression than individual genetic, CSF, PET, or structural MRI modalities [22]. However, those analyses evaluated modalities individually and did not determine how much each modality contributed within an integrated multimodal model.

Systematic leave-one-modality-out ablation provides a complementary framework for addressing this question. By removing one predefined modality at a time while holding the remaining feature set, preprocessing procedures, model architecture, and evaluation framework constant, ablation analysis quantifies the change in model discrimination associated with removal of each modality. This approach can distinguish modalities that provide substantial complementary predictive information from those whose information is largely redundant with other available features. Such analysis is particularly relevant when multimodal models require substantially more complete, expensive, or invasive data than simpler prediction strategies.

In this study, we performed a systematic multimodal ablation analysis using 2,430 participants with MCI from the Alzheimer’s Disease Neuroimaging Initiative (ADNI) [22]. A full multimodal machine-learning model incorporating clinical, cognitive, genetic, structural MRI, CSF, and PET features was used as the reference. Then five leave-one-modality-out models were evaluated by removing cognition, APOE, MRI, CSF, or PET features individually. Model discrimination was assessed using repeated stratified cross-validation, while paired out-of-fold predictions were used for formal AUC comparisons using DeLong’s test [23] with Holm-Bonferroni correction [24]. Complete-case sensitivity analyses were additionally performed to assess the potential influence of missing biomarker data [25], since not all of 2,430 participants had clinical, cognitive, genetic, structural MRI, CSF, and PET information available.

Our objective was to quantify the marginal predictive contribution of each modality within an integrated machine-learning model and to determine whether individual modalities contribute complementary information or are largely redundant with the remaining modalities, with the ultimate goal of informing low-cost clinical decision-support tools for first-line health professionals and for risk stratification in clinical trial recruitment.

## Methods

### Study Population

The study cohort comprised participants with mild cognitive impairment (MCI) at baseline drawn from the Alzheimer’s Disease Neuroimaging Initiative (ADNI) [21]. Participants were classified according to whether they progressed from MCI to Alzheimer’s disease (AD) within 24 months of baseline (progressors vs. non-progressors). The final study cohort included 2,430 participants, including 547 progressors and 1,883 non-progressors.

Baseline demographic, cognitive, genetic, neuroimaging, and fluid biomarker characteristics were summarized according to 24-month progression status. Continuous variables were reported as mean (standard deviation [SD]), and categorical variables as number (percentage). Between-group comparisons were performed using Welch’s *t* test for continuous variables and the chi-square test for categorical variables. Standardized mean differences (SMDs; Cohen’s *d*) were calculated for continuous variables. Available sample size varied by biomarker because of modality-specific missingness. Missingness ranged from less than 1% for demographic and cognitive variables to 57.9% for CSF Aβ42, reflecting differences in biomarker availability within ADNI.

### Modality Ablation Study

We conducted a systematic modality ablation study to quantify the contribution of predefined clinical, cognitive, genetic, neuroimaging, and fluid biomarker modalities to prediction of 24-month progression from mild cognitive impairment (MCI) to Alzheimer’s disease (AD). A full multimodal feature set served as the reference model, and individual modalities were removed one at a time while all remaining features were retained. This leave-one-modality-out design allowed differences in model discrimination to be attributed to the excluded modality under an otherwise identical modeling and evaluation framework.

The full multimodal model incorporated demographic variables (age and sex); cognitive assessments including the Mini-Mental State Examination (MMSE) [26], Logical Memory Delayed Recall (LDELTOTAL) [27], and Rey Auditory Verbal Learning Test Immediate Recall (RAVLT-immediate) [28]; APOE ε4 carrier status [29]; structural magnetic resonance imaging (MRI) biomarkers including hippocampal volume, entorhinal cortex volume, middle temporal gyrus volume, whole-brain volume, and ventricular volume [30]; cerebrospinal fluid (CSF) biomarkers including Aβ42, total tau, and phosphorylated tau [31]; and positron emission tomography (PET) biomarkers including AV45 amyloid PET and fluorodeoxyglucose (FDG) PET [12].

Seven prespecified feature sets were evaluated:

1. Clinical: age and sex only.
2. Multimodal: all clinical, cognitive, APOE, MRI, CSF, and PET features.
3. Multimodal without cognitive assessments: all multimodal features except MMSE, LDELTOTAL, and RAVLT-immediate.
4. Multimodal without MRI: all multimodal features except structural MRI biomarkers.
5. Multimodal without CSF: all multimodal features except CSF biomarkers.
6. Multimodal without PET: all multimodal features except PET biomarkers.
7. Multimodal without APOE: all multimodal features except APOE ε4 carrier status.

All seven models were trained and evaluated using the same study cohort, feature definitions, preprocessing procedures, and model specification. The primary prediction model was XGBoost configured for binary classification. All hyperparameters were held constant across feature sets, while the positive-class weighting parameter (scale_pos_weight) was calculated within each training fold from the ratio of non-progressors to progressors to account for class imbalance.

Missing numerical values were imputed using the median calculated from the training fold. Categorical variables were imputed using the most frequent category and subsequently one-hot encoded. Preprocessing was fitted exclusively on the training data within each cross-validation split and then applied to the corresponding validation data to prevent information leakage [32].

### Cross-Validation and Performance Evaluation

Model performance was estimated using repeated stratified 5-fold cross-validation with 10 repetitions, resulting in 50 validation folds for each model [33]. Stratification preserved the distribution of 24-month progressors and non-progressors across folds.

Within each training fold, the classification threshold was optimized using the Youden index [34] and subsequently applied to the corresponding validation fold. Performance was evaluated using the area under the receiver operating characteristic curve (AUC) [35], accuracy, balanced accuracy [36], sensitivity, specificity, precision, negative predictive value, F1 score, and Brier score [37].

AUC was the primary measure of model discrimination. Performance was summarized as the mean and standard deviation across the 50 validation folds. Ninety-five percent confidence intervals for AUC were estimated by bootstrap resampling of a prespecified out-of-fold (OOF) prediction set [38].

### Statistical Comparison of Ablation Models

Model discrimination was evaluated using two complementary AUC analyses serving distinct purposes. Primary performance estimates were obtained from repeated stratified 5-fold cross-validation with 10 repetitions, with mean AUC across the 50 validation folds reported as the principal estimate of expected discrimination. Because repeated cross-validation generates multiple validation predictions for each participant across repetitions, these predictions do not constitute a single independent OOF prediction per participant and therefore are not appropriate for DeLong comparison.

For formal pairwise comparison of model discrimination, a separate non-repeated stratified 5-fold cross-validation analysis was performed using identical fold assignments across all seven models. Within each fold, preprocessing was fitted exclusively to the training data and applied to the held-out validation data, while the same model specification and hyperparameters were maintained across feature sets. This procedure generated one paired OOF prediction per participant for each model. Pairwise AUC comparisons were performed using DeLong’s test for correlated ROC curves [23], and P values were adjusted for 21 comparisons using the Holm–Bonferroni procedure [24]. Statistical significance was defined as an adjusted P value below 0.05.

For the modality ablation analysis, the change in discrimination associated with removal of each modality was quantified relative to the full multimodal model as:

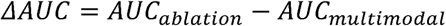

where negative values indicate reduced discrimination following removal of the specified modality.

### Complete-Case Sensitivity Analysis

Because CSF and PET biomarkers exhibited substantial missingness (Table 1), a complete-case sensitivity analysis was performed to assess the potential influence of median imputation on model discrimination.

**Table 1.**
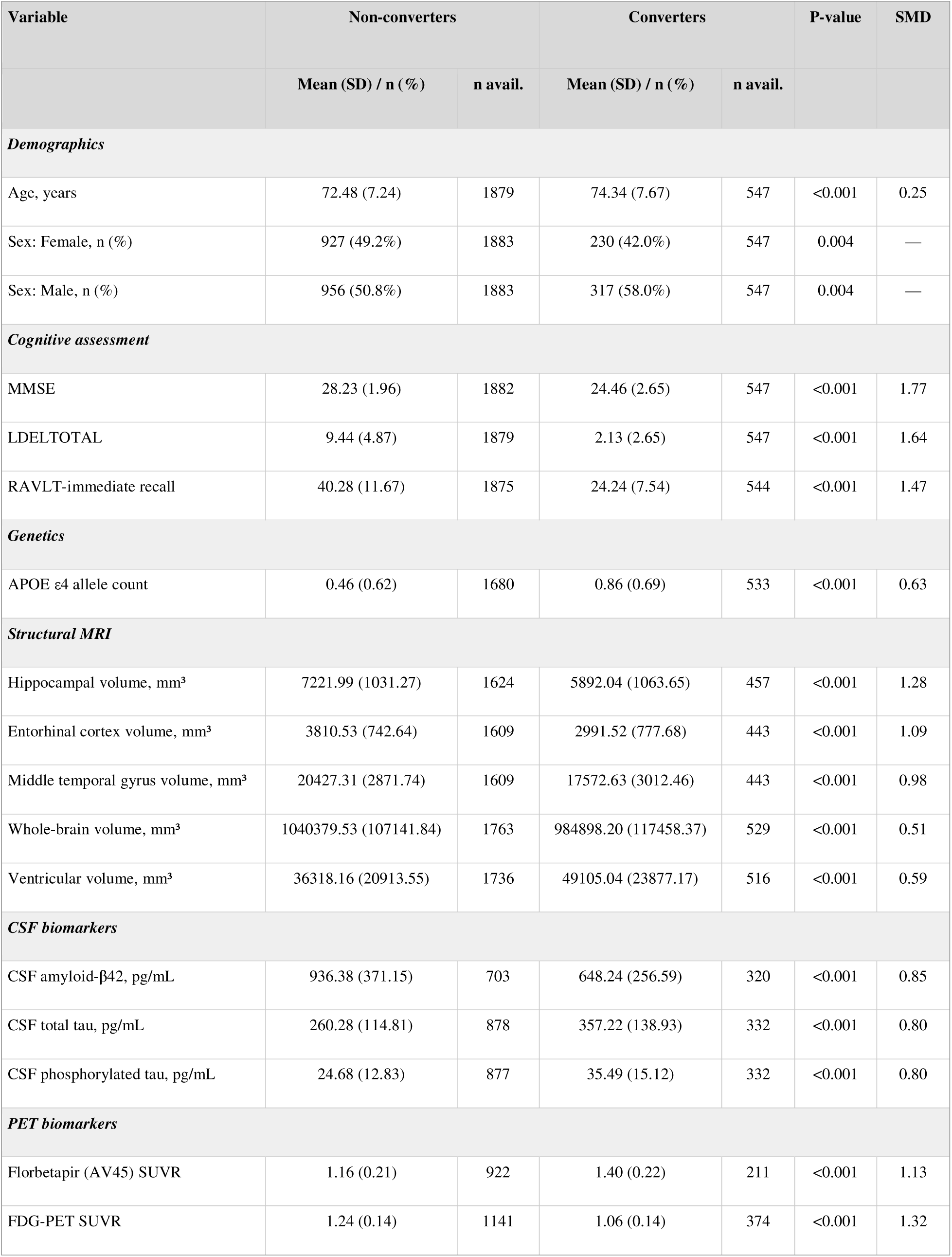
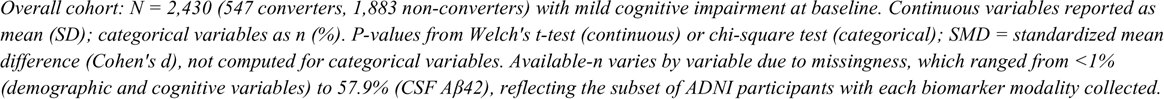
Cohort characteristics by 24-month progression status.

| Variable | Non-converters |  | Converters |  | P-value | SMD |
| --- | --- | --- | --- | --- | --- | --- |
|  | Mean (SD) / n (%) | n avail. | Mean (SD) / n (%) | n avail. |  |  |
| **Demographics** |  |  |  |  |  |  |
| Age, years | 72.48 (7.24) | 1879 | 74.34 (7.67) | 547 | <0.001 | 0.25 |
| Sex: Female, n (%) | 927 (49.2%) | 1883 | 230 (42.0%) | 547 | 0.004 | — |
| Sex: Male, n (%) | 956 (50.8%) | 1883 | 317 (58.0%) | 547 | 0.004 | — |
| **Cognitive assessment** |  |  |  |  |  |  |
| MMSE | 28.23 (1.96) | 1882 | 24.46 (2.65) | 547 | <0.001 | 1.77 |
| LDELTOTAL | 9.44 (4.87) | 1879 | 2.13 (2.65) | 547 | <0.001 | 1.64 |
| RAVLT-immediate recall | 40.28 (11.67) | 1875 | 24.24 (7.54) | 544 | <0.001 | 1.47 |
| **Genetics** |  |  |  |  |  |  |
| APOE ε4 allele count | 0.46 (0.62) | 1680 | 0.86 (0.69) | 533 | <0.001 | 0.63 |
| **Structural MRI** |  |  |  |  |  |  |
| Hippocampal volume, mm<sup>3</sup> | 7221.99 (1031.27) | 1624 | 5892.04 (1063.65) | 457 | <0.001 | 1.28 |
| Entorhinal cortex volume, mm<sup>3</sup> | 3810.53 (742.64) | 1609 | 2991.52 (777.68) | 443 | <0.001 | 1.09 |
| Middle temporal gyrus volume, mm<sup>3</sup> | 20427.31 (2871.74) | 1609 | 17572.63 (3012.46) | 443 | <0.001 | 0.98 |
| Whole-brain volume, mm<sup>3</sup> | 1040379.53 (107141.84) | 1763 | 984898.20 (117458.37) | 529 | <0.001 | 0.51 |
| Ventricular volume, mm<sup>3</sup> | 36318.16 (20913.55) | 1736 | 49105.04 (23877.17) | 516 | <0.001 | 0.59 |
| **CSF biomarkers** |  |  |  |  |  |  |
| CSF amyloid-β42, pg/mL | 936.38 (371.15) | 703 | 648.24 (256.59) | 320 | <0.001 | 0.85 |
| CSF total tau, pg/mL | 260.28 (114.81) | 878 | 357.22 (138.93) | 332 | <0.001 | 0.80 |
| CSF phosphorylated tau, pg/mL | 24.68 (12.83) | 877 | 35.49 (15.12) | 332 | <0.001 | 0.80 |
| **PET biomarkers** |  |  |  |  |  |  |
| Florbetapir (AV45) SUVR | 1.16 (0.21) | 922 | 1.40 (0.22) | 211 | <0.001 | 1.13 |
| FDG-PET SUVR | 1.24 (0.14) | 1141 | 1.06 (0.14) | 374 | <0.001 | 1.32 |

*Overall cohort: N = 2,430 (547 converters, 1,883 non-converters) with mild cognitive impairment at baseline. Continuous variables reported as mean (SD); categorical variables as n (%). P-values from Welch's t-test (continuous) or chi-square test (categorical); SMD = standardized mean difference (Cohen's d), not computed for categorical variables. Available-n varies by variable due to missingness, which ranged from <1% (demographic and cognitive variables) to 57.9% (CSF Aβ42), reflecting the subset of ADNI participants with each biomarker modality collected.*

For each feature set included in the sensitivity analysis, three complementary analyses were performed. First, the primary model was trained using fold-specific median imputation and evaluated in the full study cohort. Second, the out-of-fold (OOF) predictions from this imputed model were evaluated only among participants with complete observed values for all features included in the corresponding feature set. Third, the model was refit and evaluated exclusively on the same complete-case participants without imputation.

The latter two analyses used identical participants and outcome labels, allowing paired comparison of AUCs using DeLong’s test [23]. This sensitivity analysis was designed to determine whether training on the larger imputed cohort materially altered predictions for participants with complete biomarker data.

### SHAP-Based Feature Importance Analysis

To complement the modality-level ablation analysis with feature-level interpretability, Shapley Additive Explanations (SHAP) values [39] were calculated for the XGBoost models. SHAP values quantify the contribution of individual features to model predictions and were summarized using mean absolute SHAP values as a measure of relative feature importance.

Following completion of all cross-validation procedures, each model was refit on the complete study cohort using the same preprocessing pipeline and model specification. SHAP values were then computed using a tree-based explainer. These refitted models were used exclusively for feature-importance analysis and were not used to estimate cross-validated predictive performance or conduct statistical comparisons.

Mean absolute SHAP values were calculated for the full multimodal model and each leave-one-modality-out model. Feature importance distributions were subsequently compared across models to characterize redistribution of predictive attribution among the remaining features following removal of each biomarker modality.

## Results

### Ablation Study Results

The full multimodal model achieved the highest discrimination for 24-month progression from mild cognitive impairment (MCI) to Alzheimer’s disease, with a mean AUC of 0.934 (95% CI: 0.923–0.944), accuracy of 0.876, balanced accuracy of 0.855, sensitivity of 0.818, specificity of 0.893, and Brier score of 0.094 (Table 2).

**Table 2.** Primary performance metrics across all seven ablation models.

| Model | N Features | AUC | Accuracy | Balanced Accuracy | Sensitivity | Specificity | Brier Score |
| --- | --- | --- | --- | --- | --- | --- | --- |
| Multimodal | 16 | 0.934 (0.010) | 0.876 (0.014) | 0.855 (0.018) | 0.818 (0.039) | 0.893 (0.018) | 0.094 (0.009) |
| No APOE | 15 | 0.933 (0.010) | 0.874 (0.013) | 0.856 (0.016) | 0.823 (0.031) | 0.889 (0.017) | 0.096 (0.009) |
| No PET | 14 | 0.932 (0.011) | 0.874 (0.013) | 0.857 (0.014) | 0.826 (0.032) | 0.887 (0.018) | 0.095 (0.009) |
| No MRI | 11 | 0.932 (0.010) | 0.866 (0.017) | 0.862 (0.017) | 0.854 (0.037) | 0.869 (0.024) | 0.098 (0.010) |
| No CSF | 13 | 0.931 (0.010) | 0.873 (0.014) | 0.858 (0.016) | 0.831 (0.031) | 0.885 (0.018) | 0.096 (0.009) |
| No Cognitive | 13 | 0.883 (0.012) | 0.824 (0.016) | 0.806 (0.018) | 0.772 (0.037) | 0.840 (0.022) | 0.125 (0.008) |
| Clinical | 2 | 0.556 (0.025) | 0.539 (0.029) | 0.539 (0.025) | 0.540 (0.073) | 0.538 (0.050) | 0.244 (0.005) |

Values are mean (SD) across repeated stratified 5-fold cross-validation with 10 repetitions (50 total validation folds). Models are ordered by mean AUC. Decision thresholds were optimized within each training fold using the Youden index and subsequently applied to the corresponding validation fold. The clinical model was included as a reference model and was not considered a leave-one-modality-out ablation.

Removal of individual modalities produced markedly different effects on model discrimination. The largest reduction in AUC occurred when cognitive assessments were removed. The multimodal model without cognitive assessments achieved an AUC of 0.883, corresponding to an absolute ΔAUC of −0.051 relative to the full multimodal model. In contrast, removal of APOE, PET, MRI, or CSF produced only minimal changes in discrimination, with AUCs of 0.933, 0.932, 0.932, and 0.931, respectively. The corresponding ΔAUC values were −0.001, −0.002, −0.002, and -0.003.

The relative changes in discrimination across the leave-one-modality-out models are shown in Figure 1. Removal of cognitive assessments was clearly distinguished from the other ablations by the magnitude of the reduction in AUC.

**Figure 1.**
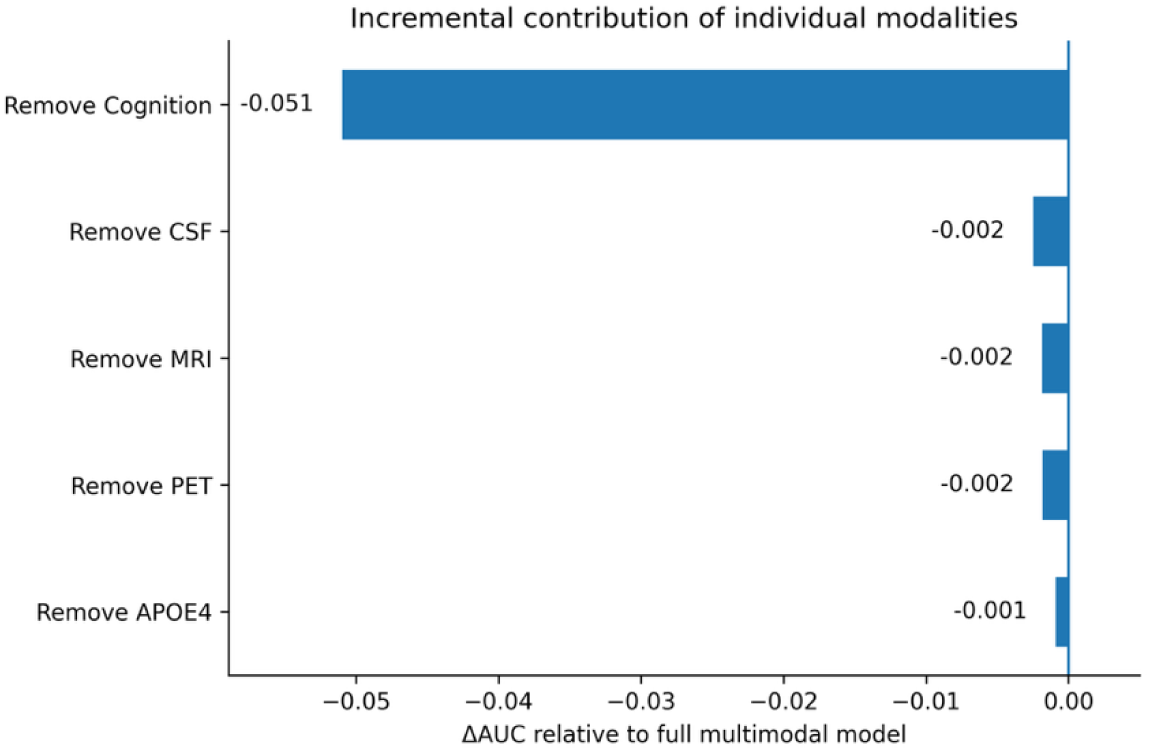
Change in AUC Relative to the Full Multimodal Model. ΔAUC for each leave-one-modality-out model relative to the full multimodal model. Negative values indicate reduced discrimination following removal of the corresponding modality. Removal of cognitive assessments produced the largest reduction in AUC (ΔAUC = −0.051), whereas removal of APOE, PET, MRI, and CSF resulted in substantially smaller changes (ΔAUC = −0.001, −0.002, −0.002, and −0.003, respectively).

The clinical model consisting of age and sex alone achieved an AUC of 0.556, indicating limited discrimination in this MCI cohort. The clinical model was included as a contextual reference for the multimodal analysis rather than as a modality-ablation comparison.

Among the leave-one-modality-out models, removal of APOE produced the smallest change in discrimination, with AUC decreasing from 0.934 to 0.933. Removal of PET and MRI each resulted in an AUC of 0.932, while removal of CSF resulted in an AUC of 0.931. These changes corresponded to reductions of no more than 0.003 AUC units relative to the full multimodal model.

Removal of MRI produced a modest increase in sensitivity from 0.818 to 0.854, accompanied by a reduction in specificity from 0.893 to 0.869. Similar modest changes in classification characteristics were observed across the other ablation models despite their relatively stable AUCs.

In contrast, removal of cognitive assessments resulted in substantially poorer overall performance. Accuracy decreased from 0.876 to 0.824, balanced accuracy from 0.855 to 0.806, and the Brier score increased from 0.094 to 0.125. Thus, cognitive assessment removal affected both discrimination and probabilistic prediction.

Overall, the ablation analysis demonstrated a pronounced asymmetry among modalities: cognitive assessments were uniquely influential within the multimodal model, whereas removal of APOE, PET, MRI, or CSF individually produced little change in discrimination.

### Statistical Comparison of Ablation Models

Pairwise DeLong comparisons were performed using single-pass, non-repeated 5-fold out-of-fold (OOF) predictions, with Holm-Bonferroni correction for the 21 pairwise comparisons among the seven models (Table 3). The clinical model differed significantly from every other model, with all comparisons remaining significant after correction (all p < 0.001). This confirms that age and sex alone provided substantially less discrimination than models incorporating cognitive and/or biomarker information.

**Table 3.** Pairwise DeLong comparisons of AUC across all seven models.

| Model A | Model B | AUC A | AUC B | ΔAUC | p-value | Holm $\alpha$ threshold | Significant (Holm) |
| --- | --- | --- | --- | --- | --- | --- | --- |
| Clinical | Multimodal | 0.545 | 0.933 | -0.389 | < 0.001 | 0.0024 | Yes |
| Clinical | No Cognitive | 0.545 | 0.885 | -0.340 | < 0.001 | 0.0025 | Yes |
| Clinical | No MRI | 0.545 | 0.930 | -0.385 | < 0.001 | 0.0026 | Yes |
| Clinical | No CSF | 0.545 | 0.931 | -0.386 | < 0.001 | 0.0028 | Yes |
| Clinical | No PET | 0.545 | 0.932 | -0.387 | < 0.001 | 0.0029 | Yes |
| Clinical | No APOE | 0.545 | 0.932 | -0.388 | < 0.001 | 0.0031 | Yes |
| Multimodal | No Cognitive | 0.933 | 0.885 | +0.049 | < 0.001 | 0.0033 | Yes |
| No Cognitive | No APOE | 0.885 | 0.932 | -0.048 | < 0.001 | 0.0036 | Yes |
| No Cognitive | No CSF | 0.885 | 0.931 | -0.046 | < 0.001 | 0.0038 | Yes |
| No Cognitive | No PET | 0.885 | 0.932 | -0.047 | < 0.001 | 0.0042 | Yes |
| No Cognitive | No MRI | 0.885 | 0.930 | -0.045 | < 0.001 | 0.0045 | Yes |
| Multimodal | No CSF | 0.933 | 0.931 | +0.002 | 0.054 | 0.0050 | No |
| Multimodal | No MRI | 0.933 | 0.930 | +0.003 | 0.087 | 0.0056 | No |
| Multimodal | No PET | 0.933 | 0.932 | +0.001 | 0.261 | 0.0063 | No |
| Multimodal | No APOE | 0.933 | 0.932 | +0.001 | 0.271 | 0.0071 | No |
| No MRI | No APOE | 0.930 | 0.932 | -0.002 | 0.279 | 0.0083 | No |
| No CSF | No APOE | 0.931 | 0.932 | -0.001 | 0.356 | 0.0100 | No |
| No MRI | No PET | 0.930 | 0.932 | -0.002 | 0.443 | 0.0125 | No |
| No CSF | No PET | 0.931 | 0.932 | -0.001 | 0.599 | 0.0167 | No |
| No MRI | No CSF | 0.930 | 0.931 | -0.001 | 0.712 | 0.0250 | No |
| No PET | No APOE | 0.932 | 0.932 | 0.000 | 0.761 | 0.0500 | No |

Among the ablation models, removal of cognitive assessments produced the only statistically significant reduction in discrimination relative to the full multimodal model. The full multimodal model achieved an OOF AUC of 0.933 compared with 0.885 for the model without cognitive assessments, corresponding to a ΔAUC of 0.049 (p < 0.001 after Holm-Bonferroni correction). In contrast, none of the differences between the full multimodal model and the other four modality-removed models reached statistical significance after correction. The OOF AUC differences were 0.001 for APOE removal, 0.001 for PET removal, 0.003 for MRI removal, and 0.002 for CSF removal.

Similarly, no statistically significant differences were observed among the models without APOE, PET, MRI, or CSF. Thus, the statistical analysis independently supported the principal ablation finding that cognitive assessments were the only modality whose removal resulted in a statistically detectable loss of discrimination.

### Complete-Case Sensitivity Analysis

A complete-case sensitivity analysis was performed to evaluate whether median imputation influenced the observed discrimination of models with substantial missingness (Table 4). For the full multimodal model, 499 participants (20.5% of the cohort) had complete data across all model features. The imputed model achieved an AUC of 0.940 when evaluated on these complete-case participants, compared with an AUC of 0.943 for a model refit exclusively on the complete-case subset. The difference was not statistically significant (p = 0.583).

**Table 4.** Complete-case sensitivity analysis: impact of median imputation on model discrimination.

| Model | N Complete<br>(% of 2,430) | AUC, Imputed<br>(All N) | AUC, Imputed Model<br>(Complete-Case<br>Subset) | AUC, Complete-Case<br>Refit (95% CI) | DeLong p<br>(Subset vs.<br>Refit) | Interpretation |
| --- | --- | --- | --- | --- | --- | --- |
| Multimodal | 499 (20.5%) | 0.933 (0.923–0.944) | 0.940 | 0.943 (0.924–0.963) | 0.583 | No significant difference |
| No CSF | 753 (31.0%) | 0.931 (0.920–0.941) | 0.942 | 0.944 (0.926–0.961) | 0.695 | No significant difference |
| No PET | 786 (32.3%) | 0.932 (0.920–0.942) | 0.936 | 0.924 (0.906–0.942) | 0.005 | Significant difference |
| No MRI | 653 (26.9%) | 0.930 (0.919–0.940) | 0.936 | 0.933 (0.913–0.952) | 0.454 | No significant difference |
| No APOE | 499 (20.5%) | 0.932 (0.922–0.943) | 0.942 | 0.944 (0.924–0.963) | 0.777 | No significant difference |
| No Cognitive | 500 (20.6%) | 0.885 (0.870–0.899) | 0.918 | 0.910 (0.882–0.936) | 0.341 | No significant difference |

*For each model with substantial missingness, three AUC estimates are compared: (1) the primary imputed model evaluated on all N=2,430 participants; (2) that same imputed model's predictions restricted to only the complete-case subset (participants with no missing values for any feature in that model); and (3) a model trained and evaluated exclusively on the complete-case subset, with no imputation. A paired DeLong test compares (2) and (3) on identical patients to isolate whether training on the larger imputed dataset changes predictions for patients who actually have complete data. Bold/shaded rows indicate a statistically significant difference ( $p < 0.05$ ).*

Similarly, no statistically significant difference between the imputed and complete-case-refit models was observed for the No CSF, No MRI, No APOE, or No Cognitive models. The No PET model was the exception. Among the 786 participants with complete data for the No PET model, the imputed model achieved an AUC of 0.936 on the complete-case subset, whereas complete-case refitting yielded an AUC of 0.924 (p = 0.005). This finding indicates that median imputation may have influenced the apparent discrimination of the No PET model and therefore warrants cautious interpretation of its small difference from the full multimodal model.

Overall, the sensitivity analysis did not alter the principal finding of the ablation analysis: removal of cognitive assessments produced the largest reduction in model discrimination.

### Receiver Operating Characteristic Analysis

Receiver operating characteristic curves based on the single-pass OOF predictions are shown in Figure 2. The full multimodal model and the models excluding APOE, PET, MRI, or CSF exhibited closely overlapping discrimination, whereas the model without cognitive assessments showed a visibly lower ROC curve. The clinical reference model showed substantially poorer discrimination than all multimodal and ablation models.

**Figure 2.**
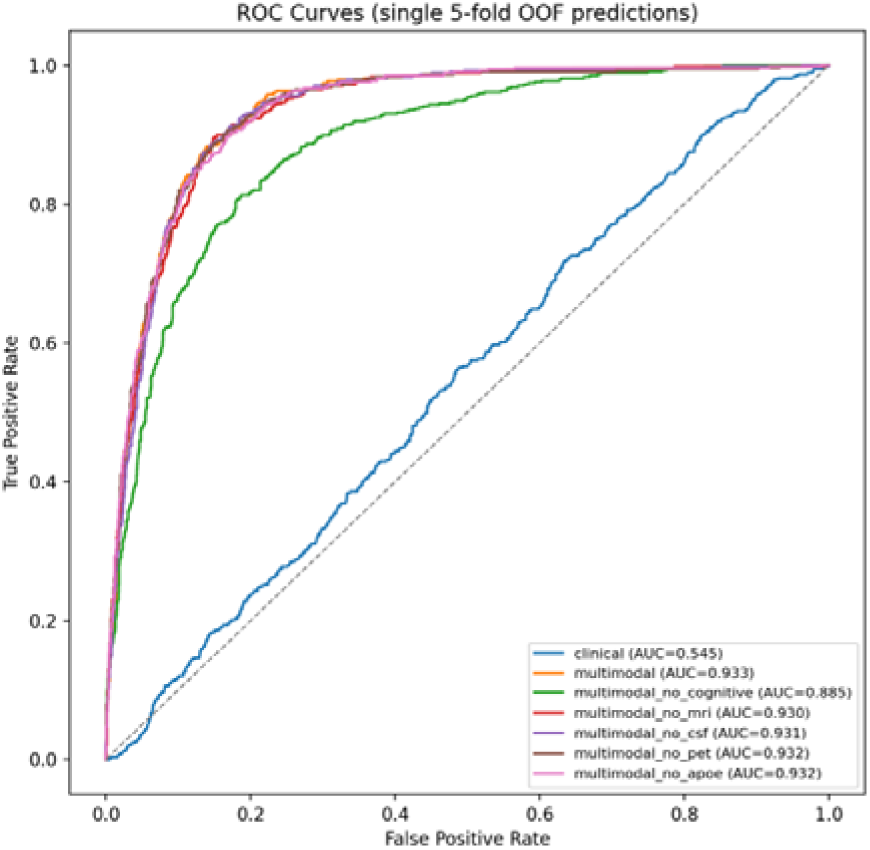
Receiver operating characteristic curves for the seven models. ROC curves for the clinical reference model, full multimodal model, and five leave-one-modality-out models using pooled single-pass OOF predictions.

**Figure 3.**
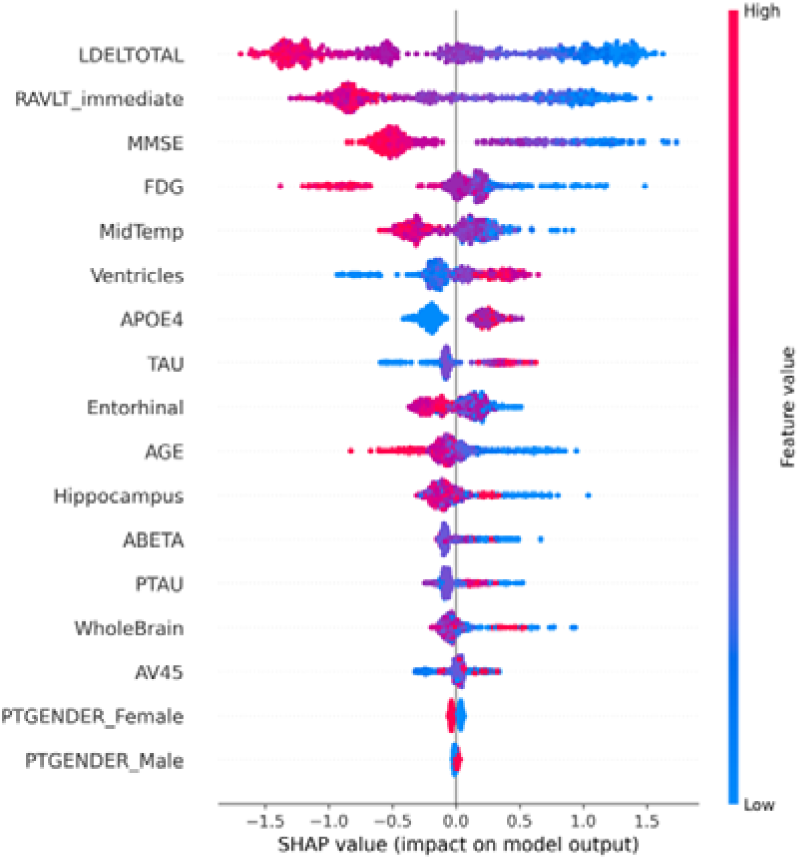
SHAP feature importance. SHAP beeswarm plot for the full multimodal model showing the distribution and direction of feature-level SHAP values across the analyzed samples.

### SHAP-Based Feature Importance

Feature-level SHAP analysis of the full multimodal model provided complementary information regarding the relative contribution of individual features to model predictions. Cognitive assessments accounted for the three highest mean absolute SHAP values. Logical Memory Delayed Recall (LDELTOTAL) was the most influential feature (mean |SHAP| = 0.894), followed by RAVLT immediate recall (0.744) and MMSE (0.599). Together, these three cognitive features represented 51.6% of the total mean absolute SHAP magnitude (Table 5). Among the non-cognitive features, FDG-PET had the largest mean absolute SHAP value (0.304), followed by middle temporal gyrus volume (0.247), ventricular volume (0.232), and APOE _ε_4 status (0.222). Other structural MRI, CSF, PET, and demographic features showed smaller individual contributions.

**Table 5.**
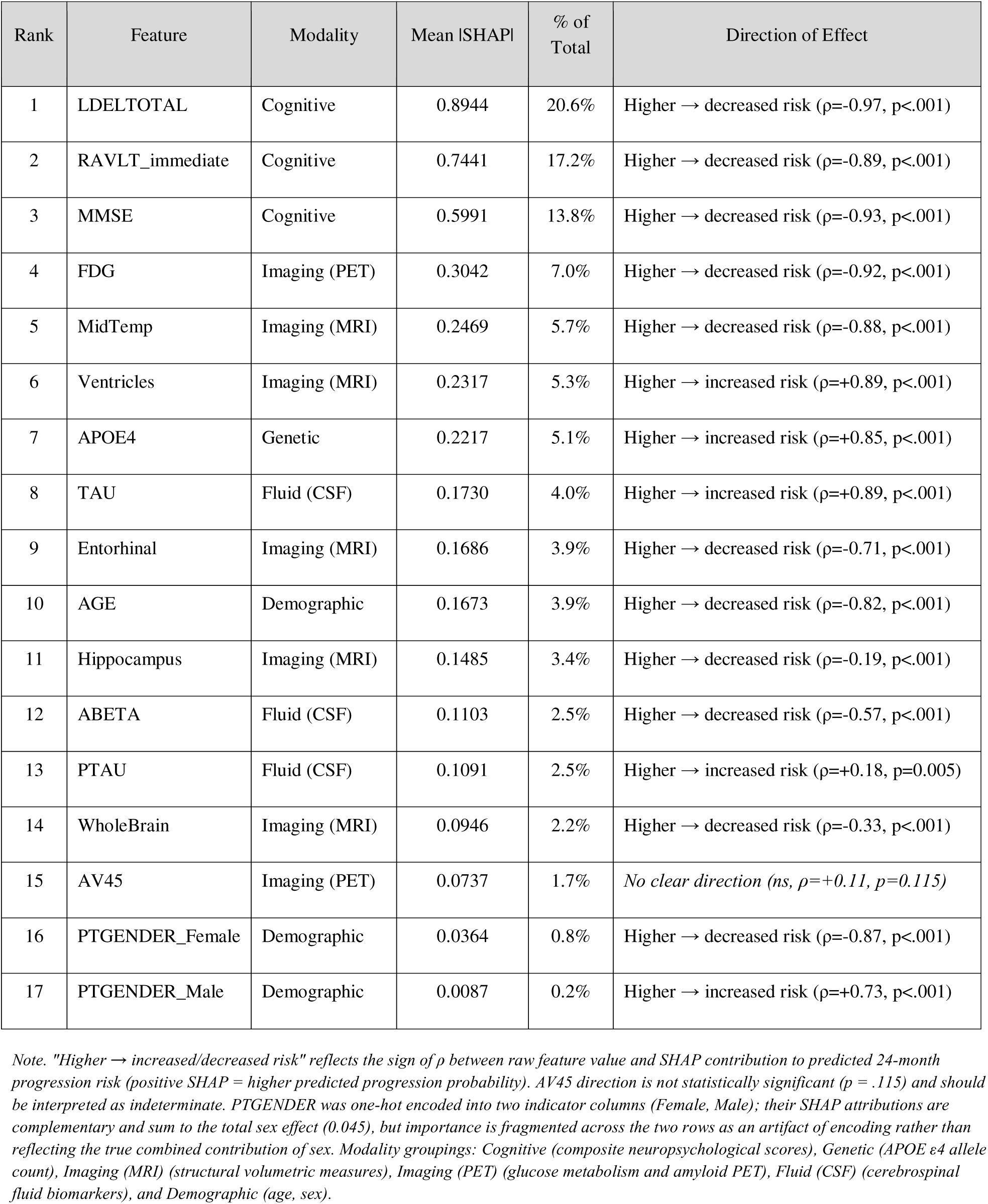
SHAP Feature Importance Ranking. Mean absolute SHAP value, relative contribution, and direction of effect for all 17 features in the full multimodal model, ranked by importance. Direction of effect is the Spearman correlation (ρ*) between each feature’s raw value and its signed SHAP contribution across n = 500 held-out test samples; missing-data biomarkers (ABETA, TAU, PTAU, AV45, FDG) use pairwise complete cases*.

The directional associations were generally consistent with established patterns of Alzheimer’s disease-related pathology. Higher cognitive scores were associated with lower predicted progression risk, while lower regional brain volumes and reduced FDG-PET metabolism were associated with higher predicted risk. Larger ventricular volume, APOE _ε_4 carrier status, and higher CSF tau measures were associated with higher predicted progression risk.

Age showed an inverse association with predicted progression risk within this MCI cohort, with higher age associated with lower predicted risk. This finding should be interpreted in the context of the selected MCI population and the prediction target rather than as a general estimate of age-related Alzheimer’s disease risk.

### SHAP Redistribution Reveals Compensatory Feature Reweighting

To examine how predictive information was redistributed following modality removal, mean absolute SHAP values were compared across the full multimodal model and the five leave-one-modality-out models (Figure 4).

**Figure 4.**
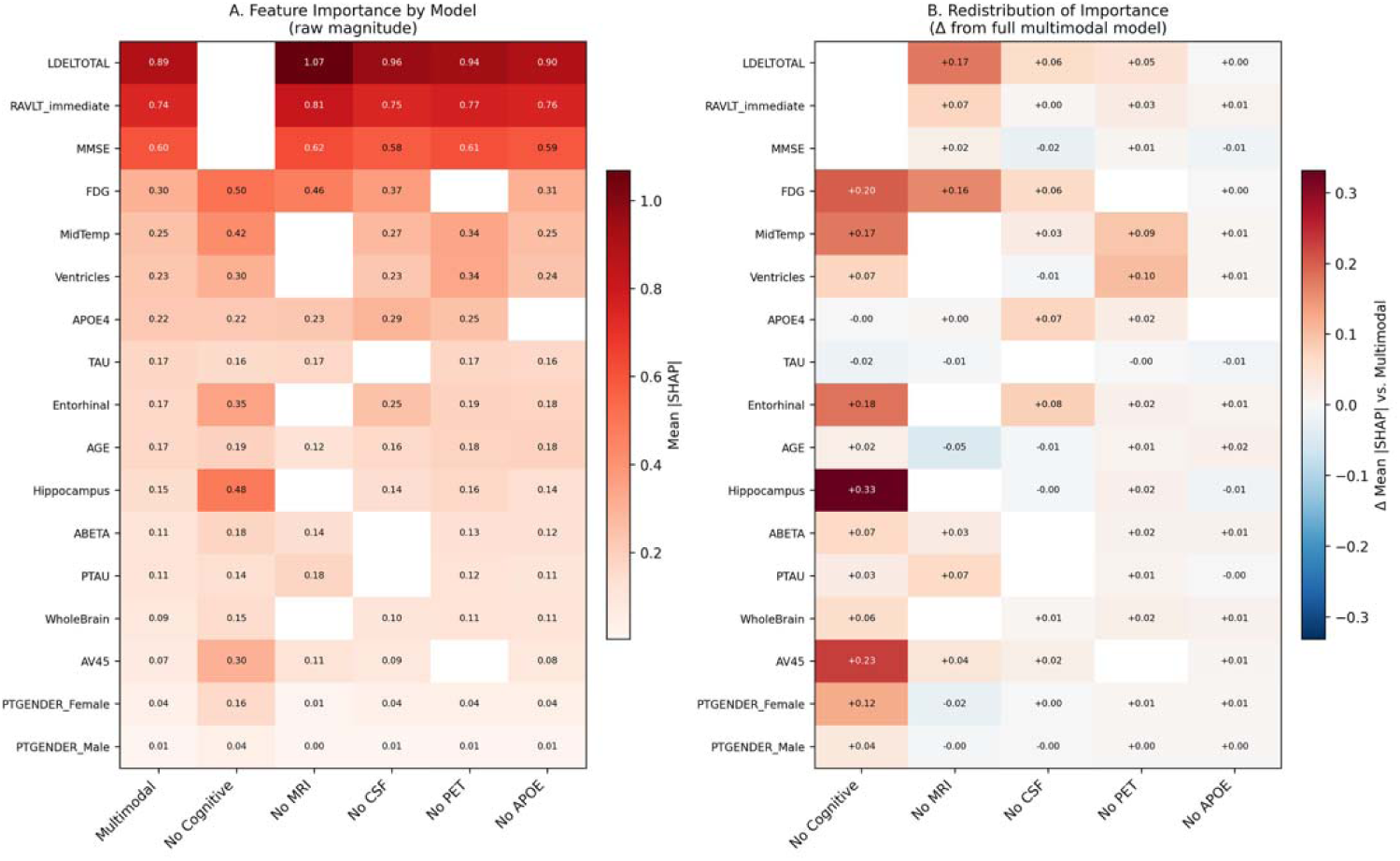
Redistribution of feature importance across ablation models. (A) Mean |SHAP| magnitude for each feature within each of the six models (full multimodal model and five leave-one-modality-out variants). (B) Change in mean |SHAP| for each feature relative to the full multimodal model when each modality is removed; positive values (red) indicate features that gained importance to compensate for the removed modality, negative values (blue) indicate features whose importance decreased. Features are ordered by baseline (multimodal) importance.

Removal of cognitive assessments produced the most pronounced redistribution of feature importance. In their absence, the model increased its reliance on remaining imaging and biomarker features. Mean |SHAP| for FDG increased from 0.304 in the full multimodal model to 0.505 following cognitive removal, while middle temporal gyrus volume increased from 0.247 to 0.421 and hippocampal volume increased from 0.149 to 0.480. AV45 increased from 0.074 to 0.303.

Removal of MRI also resulted in redistribution toward the remaining modalities. FDG-PET showed the largest proportional increase, from 0.304 to 0.463 (+52%), whereas LDELTOTAL showed the largest absolute increase, from 0.894 to 1.068 (+0.174). These changes indicate that predictive information associated with structural imaging was redistributed across both metabolic imaging and cognitive features when MRI was unavailable.

Following CSF removal, entorhinal cortex volume increased from 0.169 to 0.247, APOE ε4 from 0.222 to 0.292, and FDG-PET from 0.304 to 0.367. Following PET removal, ventricular volume increased from 0.232 to 0.335 and middle temporal gyrus volume from 0.247 to 0.339.

In contrast, removal of APOE produced little redistribution of feature importance. For example, mean |SHAP| for LDELTOTAL changed from 0.894 to 0.896, FDG from 0.304 to 0.306, and middle temporal gyrus volume from 0.247 to 0.254. This limited redistribution was consistent with the minimal change in overall discrimination following APOE removal.

Taken together, the SHAP redistribution analysis showed that removal of APOE, PET, MRI, or CSF was accompanied by reweighting of predictive importance among the remaining features, despite minimal changes in overall discrimination. In contrast, removal of cognitive assessments produced both the largest reduction in AUC and the most pronounced redistribution toward the remaining biomarker features.

## Discussion

This study evaluated the relative contribution of demographic, cognitive, genetic, structural imaging, CSF, and PET biomarkers to prediction of 24-month progression from mild cognitive impairment (MCI) to Alzheimer’s disease using a systematic modality-ablation framework. The principal finding was a pronounced hierarchy of predictive contribution: cognitive assessments accounted for the largest incremental contribution to discrimination, whereas removal of APOE genotype, structural MRI, or CSF biomarkers resulted in only small changes in model performance. PET showed a similarly small difference in the primary analysis, although its contribution was sensitive to missing-data handling. Thus, although the full multimodal model achieved the highest overall discrimination, predictive information was not distributed equally across modalities. These findings have implications for determining which biomarkers are essential, complementary, or potentially dispensable when designing clinically scalable prediction systems.

### Cognitive assessments provide the dominant predictive signal

Cognitive assessments were the most consequential modality in the ablation analysis. Removing MMSE, LDELTOTAL, and RAVLT-immediate produced the largest reduction in discrimination, with an absolute AUC decrease of approximately 0.05 from the full multimodal model. This difference remained statistically significant after correction for all pairwise comparisons, whereas none of the other individual modality-removal comparisons with the full model reached statistical significance. The finding is consistent with the established importance of episodic memory and global cognitive impairment for identifying individuals with MCI who are more likely to progress to Alzheimer’s disease dementia [6–8].

The SHAP analysis provided complementary feature-level evidence. LDELTOTAL, RAVLT-immediate, and MMSE were the three most influential individual features in the full multimodal model. Thus, the dominant contribution of cognition was not attributable to a single cognitive measure but was distributed across measures of episodic memory and global cognitive status. The convergence between modality-level ablation and feature-level importance strengthens the interpretation that cognitive assessment represents the principal source of prognostic information in this MCI cohort.

The SHAP redistribution analysis further indicated that removal of cognitive assessments did not simply eliminate predictive signal; rather, predictive importance shifted toward remaining imaging and biomarker features, including FDG-PET, hippocampal volume, middle temporal gyrus volume, and amyloid PET. This redistribution suggests that the remaining modalities contain information related to progression risk that overlaps partially with information captured by cognitive assessment. However, their increased feature importance after cognitive removal did not compensate fully for the loss of cognitive information, as reflected by the substantial decline in AUC. The results therefore support partial complementarity rather than functional equivalence among the modalities.

### Additional biomarker modalities provide limited incremental discrimination

In contrast to cognition, removal of APOE genotype, MRI, or CSF produced only small changes in discrimination relative to the full multimodal model. The absolute AUC differences were approximately 0.001-0.003, and none was statistically significant after Holm-Bonferroni correction. These findings should not be interpreted as evidence that these biomarkers lack biological relevance to Alzheimer’s disease. Rather, they indicate that their additional predictive information was limited once cognitive, demographic, genetic, imaging, and fluid features were considered jointly within this model.

This distinction between biological relevance and incremental predictive utility is important. Alzheimer’s disease biomarkers characterize partially related aspects of disease biology, including genetic susceptibility, amyloid and tau pathology, neurodegeneration, and metabolic dysfunction [14–16]. When multiple correlated manifestations of disease are simultaneously available to a prediction model, the incremental information provided by any single modality can be considerably smaller than its association with disease status when evaluated independently.

The SHAP redistribution results were consistent with this interpretation. Removal of MRI increased the contribution of both FDG-PET and cognitive features, whereas removal of CSF shifted importance toward selected imaging and APOE features. In contrast, removal of APOE produced very little redistribution of feature importance. These patterns suggest that some modalities share predictive information and that the model can reweight remaining features when one source of information is unavailable. Importantly, however, redistribution of model importance should not be interpreted as demonstrating that one biological modality biologically substitutes for another.

### PET contribution is sensitive to missing-data handling

PET warrants a more cautious interpretation because of the substantial missingness in PET measurements. In the primary analysis, removal of PET resulted in only a small reduction in AUC, and the difference from the full multimodal model was not statistically significant. However, the complete-case sensitivity analysis demonstrated a significant difference between the imputed No PET model and a model refit without imputation on the same complete-case participants. This pattern was not observed for the other modality-removal analyses examined in the sensitivity analysis.

These findings do not establish that PET is more or less important than MRI, CSF, or APOE. Instead, they demonstrate that the estimated incremental contribution of PET is particularly sensitive to the treatment of missing data. Median imputation can preserve sample size but may also alter the effective information structure of a modality with substantial missingness [25]. Consequently, the small AUC difference observed in the primary analysis should not be interpreted as definitive evidence that PET provides negligible incremental information. The appropriate conclusion is that its incremental contribution could not be estimated with the same confidence as that of modalities with more complete data. Studies using cohorts with more systematically acquired PET measurements will be important for resolving this uncertainty.

### Demographic variables alone provide limited discrimination within an MCI cohort

The clinical model containing only age and sex demonstrated near-chance discrimination (AUC 0.556). This finding illustrates the distinction between population-level risk factors and individual-level short-term prognostic discrimination. Age is a well-established risk factor for Alzheimer’s disease at the population level, but among individuals who have already entered an MCI cohort, age and sex alone provide limited information for distinguishing those who will progress to Alzheimer’s disease within 24 months.

The SHAP analysis identified an inverse association between age and predicted progression risk within this particular cohort, with younger participants receiving higher predicted risk. This finding should not be interpreted as evidence that younger age is intrinsically associated with greater Alzheimer’s disease risk. Rather, it may reflect the composition of an MCI-selected research cohort, in which younger individuals with MCI who subsequently progress may represent a subgroup with different underlying pathological or clinical characteristics, while older individuals who remain stable may include patients with slower-progressing or non-Alzheimer’s causes of cognitive impairment. Because this association is observational and cohort-specific, it should be considered a model-derived pattern requiring replication rather than a general epidemiological conclusion.

### Redistribution of predictive information across modalities

The SHAP redistribution analysis provided additional insight into the ablation results. Removal of individual modalities generally resulted in increased importance of selected features from the remaining modalities, suggesting that the model retained partially overlapping sources of predictive information. This pattern was particularly evident following removal of MRI, CSF, and PET, where importance shifted toward combinations of cognitive, structural, metabolic, and genetic features. In contrast, removal of APOE produced minimal redistribution, consistent with its very small effect on overall discrimination. The most pronounced redistribution occurred after removal of cognitive assessments, with greater reliance on imaging and other biomarker features; however, this compensatory reweighting did not prevent a substantial reduction in AUC. These findings suggest that the remaining modalities can partially compensate for the absence of one another, but the predictive information carried by cognitive assessment is less readily recoverable from the other modalities.

### Implications for biomarker selection and clinical implementation

These findings suggest that cognitive assessment may serve as an effective first-line risk stratification tool in resource-limited clinical settings. Although the full multimodal model achieved the highest discrimination, removing MRI, CSF, PET, or APOE resulted in only modest reductions in performance, indicating that much of the predictive information is captured by routinely collected cognitive measures. This creates the opportunity to develop inexpensive, non-invasive screening tools that reduce dependence on costly or less accessible biomarker testing.

The results also have implications for clinical trial recruitment. A cognition-based machine learning screening layer could identify individuals at elevated risk of progression before referral for MRI, PET, or CSF confirmation, potentially reducing the number of expensive biomarker assessments required during participant screening. Rather than replacing biological biomarkers, cognitive assessment may provide a scalable and cost-effective triage strategy that prioritizes patients for more comprehensive evaluation.

### Limitations

Several limitations should be considered when interpreting these findings.

First, the analysis was conducted in a single research cohort from ADNI, and modality contributions may differ in independent populations with different demographic characteristics, disease spectra, clinical practices, and biomarker acquisition patterns. External validation is therefore necessary before the observed hierarchy of predictive contribution can be generalized to routine clinical populations.

Second, the ablation framework evaluated modality-level removal rather than individual biomarker-level removal within each modality. Consequently, the analysis establishes the incremental contribution of predefined modality groups but does not determine whether particular MRI, CSF, or PET biomarkers within those groups are individually dispensable. Future studies should evaluate within-modality ablation and identify smaller biomarker sets that preserve predictive performance.

Third, the ablation results are conditional on the feature set, prediction horizon, preprocessing strategy, and machine-learning model used in this study. A modality that provides little incremental information in the present 24-month prediction task could have greater value for longer-term prediction, different disease stages, or alternative clinical endpoints.

Fourth, SHAP values characterize model behavior rather than causal biological mechanisms. Redistribution of feature importance should therefore be interpreted as statistical reweighting within the predictive model, not as evidence that one biological modality substitutes for another. Likewise, limited incremental predictive value does not imply limited biological importance. The present analysis evaluates predictive utility rather than biological causality.

A further limitation concerns statistical comparison of AUCs. The primary performance estimates were derived from repeated stratified cross-validation, whereas formal DeLong comparisons were performed using a separate single non-repeated 5-fold cross-validation pass to obtain one paired out-of-fold prediction per participant. This approach avoids treating correlated repeated cross-validation predictions as independent observations but means that the inferential analysis and primary performance estimation are based on different cross-validation procedures. The single-pass analysis may also have limited power to detect very small differences between highly similar models.

Finally, the interpretation of PET is constrained by substantial missingness and sensitivity to missing-data handling. The complete-case sensitivity analysis indicates that the primary imputed analysis may not fully characterize PET’s incremental contribution. Additional validation in cohorts with more complete and systematically acquired PET measurements will be necessary to determine whether this finding generalizes.

## Conclusion

In this systematic modality-ablation analysis of 24-month MCI-to-Alzheimer’s progression, cognitive assessment was the dominant source of predictive information: its removal was the only modality- removal comparison to significantly reduce discrimination, whereas removing APOE, MRI, or CSF produced only small, statistically indistinguishable changes.

External validation in independent, more diverse cohorts is needed before this hierarchy can inform practice.

This finding has a direct clinical implication in primary care settings and clinical trials. Because simple, low-cost, pencil-and-paper cognitive assessments capture most of the achievable discrimination, Primary Care Physicians may be able to stratify MCI patients by risk of near-term progression using tools already available at the point of care to prioritize referral to specialists. The same low-cost stratification could support enrollment screening for clinical trials, identifying likely progressors before committing patients to more invasive or expensive confirmatory testing.

## List of Abbreviations

Aβ42: Amyloid-beta 42
AD: Alzheimer’s disease
ADNI: Alzheimer’s Disease Neuroimaging Initiative
APOE: Apolipoprotein E
AUC: Area under the receiver operating characteristic curve
CI: Confidence interval
CSF: Cerebrospinal fluid
FDG: Fluorodeoxyglucose
MCI: Mild cognitive impairment
MMSE: Mini-Mental State Examination
MRI: Magnetic resonance imaging
PET: Positron emission tomography
RAVLT: Rey Auditory Verbal Learning Test
ROC: Receiver operating characteristic

## Declarations

### Ethics approval and consent to participate

This study involved a secondary analysis of de-identified data obtained from the Alzheimer’s Disease Neuroimaging Initiative (ADNI) database. All ADNI participants provided written informed consent at their respective participating institutions, and the ADNI study received approval from the Institutional Review Board (IRB) at each participating site in accordance with the Declaration of Helsinki and applicable regulatory requirements. The present secondary analysis used publicly available de-identified data and did not involve direct contact with human participants.

## Consent for publication

Not applicable.

## Availability of data and materials

The data analyzed during the current study are available from the Alzheimer’s Disease Neuroimaging Initiative (ADNI) repository to qualified investigators upon application and approval. Information on data access is available at https://adni.loni.usc.edu. The code used to perform the analyses described in this study is available from the corresponding author upon reasonable request.

## AI Use Disclosure

The author used ChatGPT (OpenAI) and Claude (Anthropic) to assist with manuscript drafting, language editing, and reference/citation verification. The AI tool was not used for study design, data analysis, statistical computation, or interpretation of results, and the author reviewed, verified, and takes full responsibility for all content in this manuscript.

## Data Availability

The data analyzed in this study are available from the Alzheimer's Disease Neuroimaging Initiative (ADNI) repository (adni.loni.usc.edu ) upon application and approval. The derived machine-learning models, analysis code, and supplementary materials are available in the accompanying MedRxiv preprint and/or from the corresponding author upon reasonable request.

https://adni.loni.usc.edu

## Acknowledgements

Data collection and sharing for this project were supported by the Alzheimer’s Disease Neuroimaging Initiative (ADNI) (National Institutes of Health Grant U01 AG024904) and the Department of Defense ADNI (Award Number W81XWH-12-2-0012). ADNI is funded by the National Institute on Aging, the National Institute of Biomedical Imaging and Bioengineering, and through contributions from numerous public and private organizations. The grantee organization is the Northern California Institute for Research and Education, and the study is coordinated by the Alzheimer’s Therapeutic Research Institute at the University of Southern California. ADNI data are disseminated by the Laboratory for Neuro Imaging at the University of Southern California.

